# Racial and ethnic disparities in maternal mental health during COVID-19

**DOI:** 10.1101/2021.11.30.21265428

**Authors:** Ananya S. Iyengar, Tsachi Ein-Dor, Emily X. Zhang, Sabrina J. Chan, Anjali J. Kaimal, Sharon Dekel

## Abstract

Knowledge of childbirth outcomes of Black and Latinx individuals during the coronavirus pandemic is limited. Black/African American and Latinx/Hispanic individuals were matched to non-Hispanic white individuals on socio-demographics. Minority individuals were nearly three times more likely to have clinically significant traumatic stress in response to childbirth and two times more likely to report postpartum depression. Unplanned Cesarean rates were higher and incidences of skin-to-skin and breastfeeding were lower in the minority group. Racial and ethnic maternal disparities exist during COVID-19.

## Introduction

On March 11, 2020, the World Health Organization declared the rapid spread of the novel coronavirus (COVID-19) outbreak a pandemic. Peripartum individuals were subject to unique health concerns and interruptions in perinatal care (Mayopoulos et al., 2021a). Although rise in maternal morbidity has been documented among people who had and did not have COVID-19 (Chmielewska et al., 2021, Mayopoulos et al., 2021b), knowledge on peripartum adaptation of Black and Latinx individuals is limited.

Differences in traditional obstetric outcomes are well documented in pre-pandemic samples. Black and Latinx individuals have increased risk for maternal mortality and life-threatening obstetrical complications (Gyamfi-Bannerman et al., 2020). These disparities are thought to be due to systemic racism and social determinants of health. Gaps in maternal mental health outcomes are also noted. Black and Latinx individuals are more likely to report peripartum depression (Howell et al., 2005), which can exert adverse outcomes for the developing infant, impair the formation of maternal-infant attachment, and reduce maternal quality of life. Limited recognition has been given to trauma-related psychopathology triggered by childbirth (Dekel et al., 2019). Although racial and ethnic gaps exist in trauma exposure and trauma pathology (Roberts et al., 2011), whether they extent to the birth experience and the postpartum period remains unknown.

We studied individuals who gave birth during COVID-19. Using propensity score matching (PSM), we matched those identified as either Black or Latinx with the group identified as non-Hispanic white on sociodemographic factors in order to examine the impact of COVID-19 on childbirth-related outcomes of minority individuals.

## Methods

### Participants

This study is part of a project on maternal wellness launched on April 2nd, 2020 (see Mayopoulos et al., 2021b for information). Women who had given birth in the last six months, approached mainly via hospital announcements and social media, completed an anonymous survey about childbirth and mental health. Mass General Brigham Human Research Committee granted the study exemption. Here, we report on a sample of 236 women identified as either Black/African American or Hispanic/Latinx (“minority group”) and 236 matched individuals identified as non-Hispanic white.

Participants were on average two months postpartum, 31 years old, resided in the U.S. (83.7%), middle-class income (54.2%), married (78.2%), had at least college degree (65.1%), delivered a healthy baby at term (95.6%) and were primiparas (53.2%).

### Measures

Postpartum depression was measured with the Edinburgh Postnatal Depression Scale (EPDS) (Cox et al., 1987), which is used in routine care to screen for depression, and defined with a cutoff >=12 (α = 0.89).

Acute traumatic stress response to childbirth was measured with the Peritraumatic Distress Inventory (PDI) (Brunet et al., 2001). It measures emotional/physiological experiences during and shortly after a traumatic event (here, childbirth). A cutoff >=17 determines clinically significant symptoms (α = 0.86).

PTSD in relation to childbirth (CB-PTSD) was measured with the PTSD Checklist for DSM-5 (PCL-5). It is the standard self-report of PTSD symptoms regarding a specified event (here, childbirth). A cutoff >= 33 is indicative of provisional PTSD diagnosis (α = 0.91). History of sexual/physical abuse was assessed with items on the Life Events Checklist for DSM-5 (LEC-5).

Information concerning gestational week, obstetrical complications, induction, delivery mode, pregnancy complications and mental health history, neonatal intensive care unit (NICU) admission, skin-to-skin, rooming-in, breastfeeding, and demographics was collected using single items.

### Data Analysis

To create matched groups with similar background characteristics between Black or Latinx and non-Hispanic white individuals, we conducted a PSM procedure using the *MatchIt* R package. Groups were matched on maternal age and marital, employment, education, and income status, and residence country, month postpartum, and survey completion date. The estimation algorithm was logistic regression, and the matching algorithm was nearest neighbor matching with caliper of 0.2.

Following the matching, we performed a series of chi-square tests for independence of measures with Fisher’s exact test to estimate group differences in prior trauma, mental health and pregnancies, and recent childbirth outcomes (Table 1); and series of logistic regressions to examine differences in mental health outcomes controlling for mental health and abuse history, and pregnancy complications, and complications in recent childbirth (i.e., unplanned Cesarean, obstetrical complications, and NICU). We conducted a mediation analysis using R *mediation* package to examine the relationship between minority status and birth-related acute stress and associated CB-PTSD. Significance was estimated by bias-corrected bootstrap analysis with 1,000 resampling cycles (Figure 1).

**Table 1.**
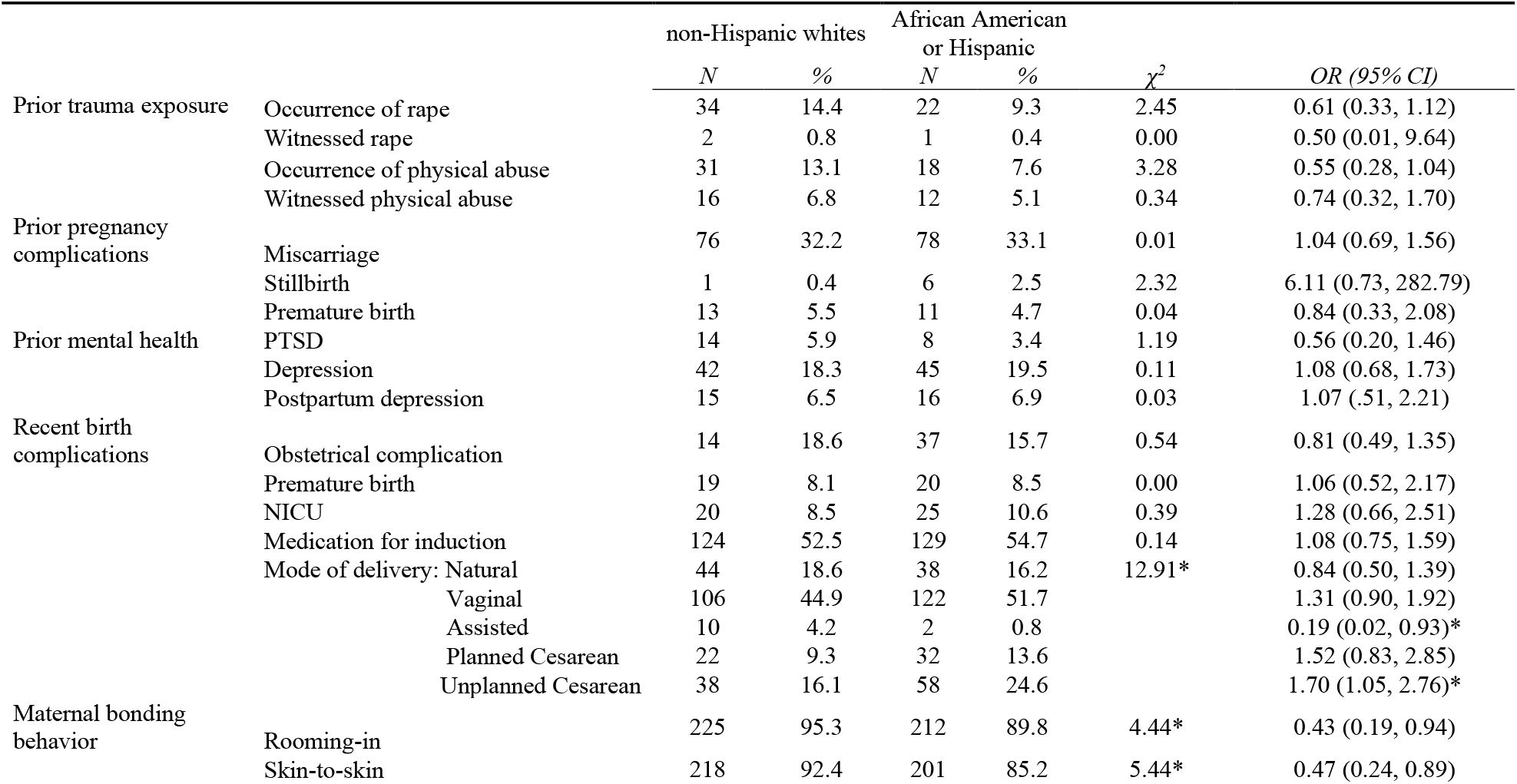
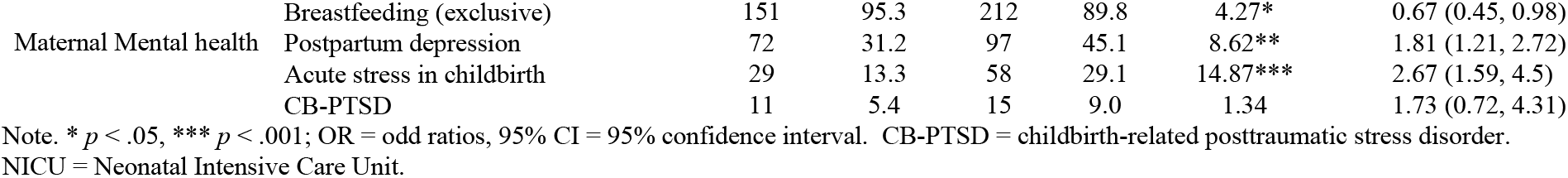
Differences in the rates of pregnancy and background factors as a function of under-represented ethnic and racial minority group

**Figure 1.**
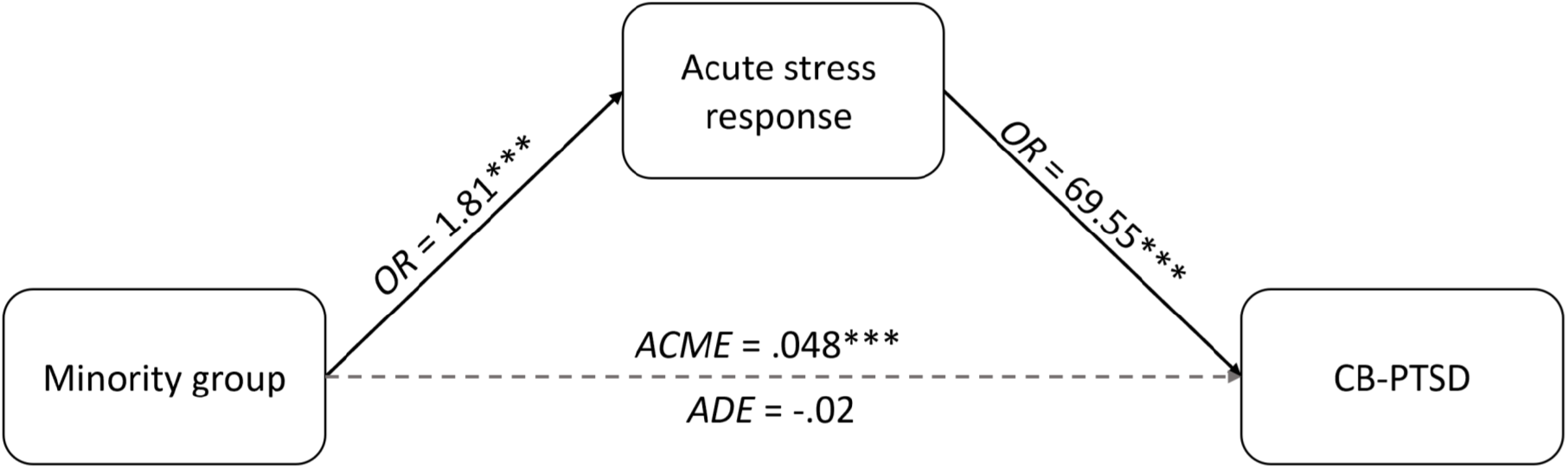
Acute childbirth-related traumatic stress response significantly mediates the effect of minority group on the likelihood of developing childbirth-related posttraumatic stress disorder (CB-PTSD). *OR* = odds ratio, *ACME* = average causal mediated effect, *ADE* = average direct effect.

## Results

The minority group were close to two times more likely to report probable postpartum depression in comparison to matched Non-Hispanic white individuals, although there were no differences between the groups in depression history. The minority group was close to three times more likely to report clinically significant traumatic stress response to childbirth and higher rates of unplanned Cesareans (and lower rates of vaginal assisted), although there were no group differences in rates of history of pregnancy complications or abuse and PTSD before childbirth. The minority group had fewer incidences of immediate skin-to-skin contact, rooming-in, and exclusive breastfeeding.

Group differences in postpartum depression and acute stress remained significant after controlling for the effects of history of mental health, abuse, and complications in recent childbirth – *OR* (95% CI) = 2.9 (1.69, 5.10), *p* < .001 for acute stress response, and *OR* (95% CI) = 1.97 (1,29, 3.04) for PPD [Aside from minority status, unplanned Cesarean was a strong predictor of mental health outcome: *OR* = 2.81 (1.60, 5.00), *p* < .0001 for PPD and *OR* = 4.1 (2.23, 7.58), *p* < .0001 for acute stress].

Among minority women, having acute traumatic stress to childbirth was strongly linked with CB-PTSD – only 0.7% of women without acute stress reported CB-PTSD as compared with 28.2% of women with acute stress, *χ*^*2*^_(1)_ = 69.55, *p* = 2.2^-16^, *OR* (95% CI) = 54.79 (12.69, 497.03). The mediation path (i.e. indirect path) from minority women to CB-PTSD via acute stress response was, therefore, significant, (*ACME* (average causal mediated effect) = .048, 95% CI = .02, .08, *p* = 2.0^-16^), whereas the direct path (minority to CB-PTSD controlling for acute stress) was not, *ADE* (average direct effect) = -.02, 95% CI = -.06, .03, *p* = .58 (Figure 1).

## Discussion

Negative pregnancy outcomes represent not only a personal but also a societal burden, because they are associated with enduring health complications and costs for the mother and the developing infant. They can increase the risk of complications in subsequent pregnancies and in extreme cases lead to maternal death.

We report that Black and Latinx individuals who gave birth during COVID-19 and are comparable on important sociodemographic factors– which are known to strongly influence pregnancy and birth outcomes– to non-Hispanic white individuals nevertheless have a heavier burden of maternal psychiatric morbidity. The disproportionate negative mental health outcomes among minority individuals remains even after accounting for the influence of pre-morbid mental health, history of abuse, and complications in recent delivery. The minority women are also more likely to have unplanned Cesarean deliveries, which can carry greater risk for maternal morbidity, and are less likely to engage in immediate postpartum bonding behaviors such as skin-to-skin and breastfeeding during the sensitive period for the formation of maternal-infant attachment that is instrumental for healthy child development. This implies vast and concerning ethnic and racial disparities in maternal and infant health.

We found that Black and Latinx individuals have a high risk for experiencing their childbirth as traumatic. Approximately one third of them reported clinically significant acute stress in response to childbirth, and their risk was close to three times more than non-Hispanic white individuals. This level of distress was strongly indicative of CB-PTSD symptoms at a level of provisional diagnosis of PTSD. We also found that Black and Latinx individuals are nearly two times more likely to report postpartum depression symptoms at a level of probable depressive disorder which warrants clinical assessment.

Structural inequities and racism are the basis for health inequities and may contribute to negative maternal outcomes by functioning as psychosocial stressors (Alhusen et al., 2016). While social and economic disadvantages have been shown to result in differential access to healthcare systems, adjustment for sociodemographic factors has not fully explained gaps in pregnancy-related morbidity (Howell, 2018). Accordingly, we observe gaps in maternal mental health and maternal bonding behaviors during the pandemic not explained by social determinants of health such as income, education, geographic location and abuse history. We further document that the gaps are not accounted for by prior mental health or complications in recent childbirth. Hence, differential exposure to stressors that may reduce adaptive coping may not adequately account for negative maternal outcomes. This points to other pathways possibly at the level of the healthcare system by which racism impacts birth outcomes.

Shortcomings include reliance on self-reports that enabled performing a study swiftly during the heights of the pandemic but no inclusion of medical records and diagnostic-assessments. The Internet-based sample may introduce sample selection.

In conclusion, we demonstrate elevated mental health morbidity following childbirth in Black and Latino individuals who gave birth during COVID-19. Major efforts are needed to support successful postpartum adjustment of minority individuals through improvements in clinical practice and public policy.

## Data Availability

All data produced in the present work are contained in the manuscript.

## Notes

### Competing Interest Statement

The authors have declared no competing interest.

### Funding Statement

Sharon Dekel was supported by awards from the Eunice Kennedy Shriver National Institute of Child Health and Human Development (R21HD100817; R03HD101724) and an award from the MGH Executive Committee on Research (ISF award).

### Author Declarations

Mass General Brigham Human Research Committee granted study exemption.

## References

Alhusen, J.L., Bower, K.M., Epstein, E., Sharps, P., 2016. Racial Discrimination and Adverse Birth Outcomes: An Integrative Review. Journal of Midwifery Womens Health 61, 707–720.

Brunet, A., Weiss, D.S., Metzler, T.J., Best, S.R., Neylan, T.C., Rogers, C., Fagan, J., Marmar, C.R., 2001. The Peritraumatic Distress Inventory: a proposed measure of PTSD criterion A2. American Journal of Psychiatry 158, 1480–1485.

Chmielewska, B., Barratt, I., Townsend, R., Kalafat, E., van der Meulen, J., Gurol-Urganci, I., O’Brien, P., Morris, E., Draycott, T., Thangaratinam, S., Le Doare, K., Ladhani, S., von Dadelszen, P., Magee, L., Khalil, A., 2021. Effects of the COVID-19 pandemic on maternal and perinatal outcomes: a systematic review and meta-analysis. Lancet Global Health 9, e759–e772.

Cox, J.L., Holden, J.M., Sagovsky, R., 1987. Detection of postnatal depression: development of the 10-item Edinburgh Postnatal Depression Scale. The British Journal of Psychiatry 150, 782–786.

Dekel, S., Thiel, F., Dishy, G., Ashenfarb, A.L., 2019. Is childbirth-induced PTSD associated with low maternal attachment? Archives of Womens Mental Health 22, 119–122.

Dekel, S., Ein-Dor T., Dishy A., Mayopoulos, G.A., 2020. Beyond postpartum depression: posttraumatic stress-depressive response following childbirth. Archives of Womens Mental Health 23, 557–564.

Gyamfi-Bannerman, C., Pandita, A., Miller, E.C., Boehme, A.K., Wright, J.D., Siddiq, Z., D’Alton, M.E., Friedman, A.M., 2020. Preeclampsia outcomes at delivery and race. Journal of Maternl and Fetal Neonatal Medicine 33, 3619–3626.

Howell, E.A., Mora P.A., Horowitz, C.R., Leventhal H., 2005. Racial and ethnic differences in factors associated with early postpartum depressive symptoms. Obstetrics and Genecology 105, 1142–1150.

Howell, E.A., 2018. Reducing Disparities in Severe Maternal Morbidity and Mortality. Clinical obstetrics and gynecology 61, 387–399.

Mayopoulos, G.A., Ein-Dor, T., Li, K.G., Chan, S.J., Dekel, S., 2021a. COVID-19 positivity associated with traumatic stress response to childbirth and no visitors and infant separation in the hospital. Scientific Reports 11, 13535.

Mayopoulos, G.A., Ein-Dor, T., Dishy, G.A., Nandru, R., Chan, S.J., Hanley, L.E., Kaimal, A.J., Dekel, S., 2021b. COVID-19 is associated with traumatic childbirth and subsequent mother-infant bonding problems. Journal of Affective Disorders 282, 122–125.

Roberts A.L., Gilman, S.E., Breslau J., Breslau N, Koenen KC., 2011. Race/ethnic differences in exposure to traumatic events, development of post-traumatic stress disorder, and treatment-seeking for post-traumatic stress disorder in the United States. Psychological Medicine 41, 71–83.

Thomas, J.L., Carter, S.E., Dunkel Schetter, C., Sumner, J.A., 2021. Racial and ethnic disparities in posttraumatic psychopathology among postpartum women. Journal of Psychiatric Research 137, 36–40.

